## Supplementary material for "Spatio-temporal modelling of the first Chikungunya epidemic in an intra-urban setting: the role of socioeconomic status, environment and temperature": S1 Appendix

| | | WAIC |
|---|---|---|
| Model 0 | $\log(\mu_{i,t}) = \log(e_i) + \beta_0 + \phi_i$ | 34114.6 |
| Model 1 | $\log(\mu_{i,t}) = \log(e_i) + \beta_0 + X'_i \beta_{k,t}$ | 26418.3 |
| Model 2 | $\log(\mu_{i,t}) = \log(e_i) + \beta_0 + X'_i \beta_{k,t} + \phi_i$ | 19656.6 |
| Model 3 | $\log(\mu_{i,t}) = \log(e_i) + \beta_0 + U_{i,t} + \phi_i$<br>$U_{i,t} = \rho_i U_{i,t-1} + \zeta_i Temperature_{i,t}$ | * |
| Model 4 | $\log(\mu_{i,t}) = \log(e_i) + \beta_0 + X'_i \beta_{k,t} + U_{i,t} + \phi_i$<br>$U_{i,t} = \rho_i U_{i,t-1} + \zeta_i Temperature_{i,t}$ | 17934.8 |

* Model 3 showed lack of convergence.
