## Supplementary figures and images for "Spatio-temporal modelling of the first Chikungunya epidemic in an intra-urban setting: the role of socioeconomic status, environment and temperature"

### S1 Fig

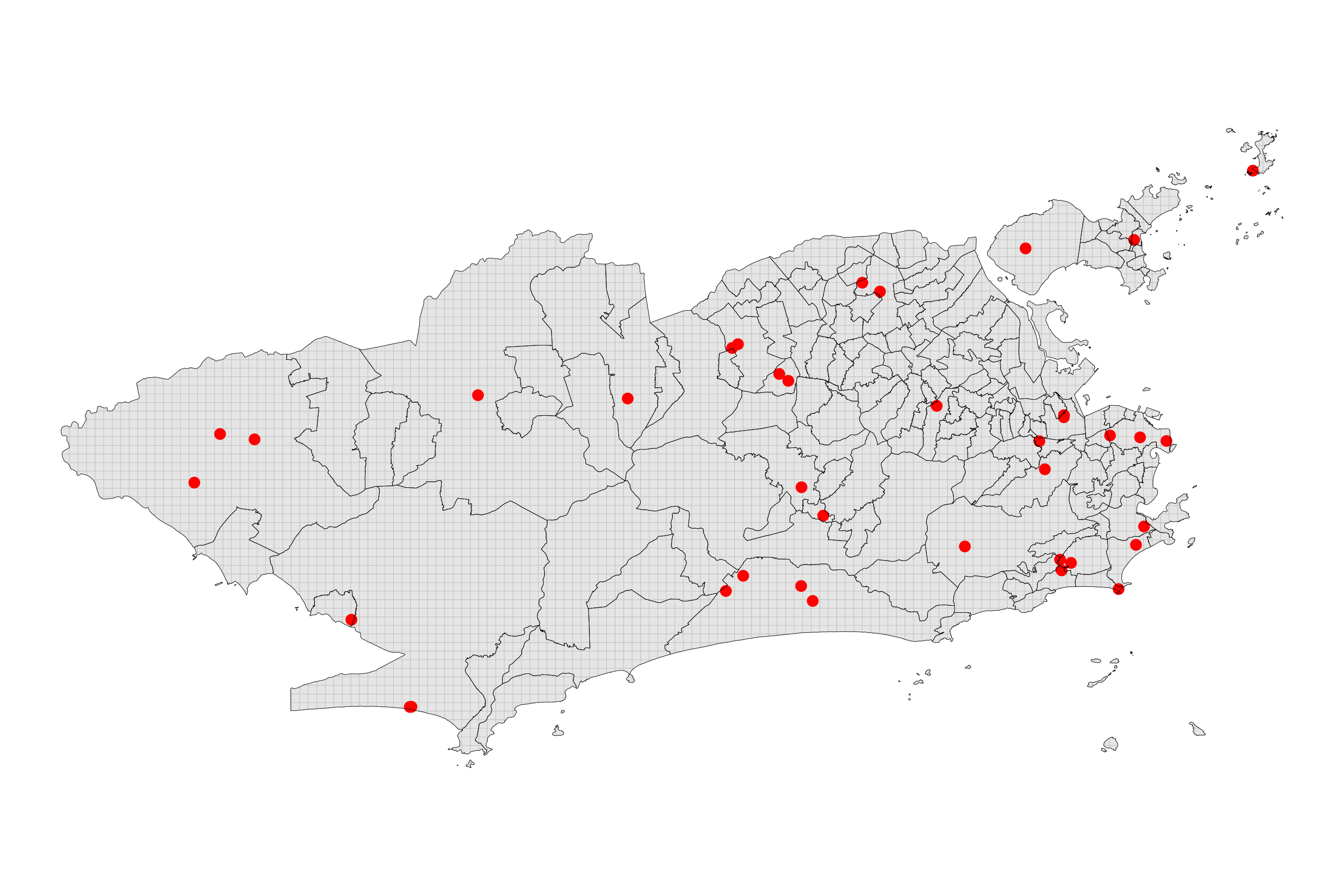

### S2 Fig

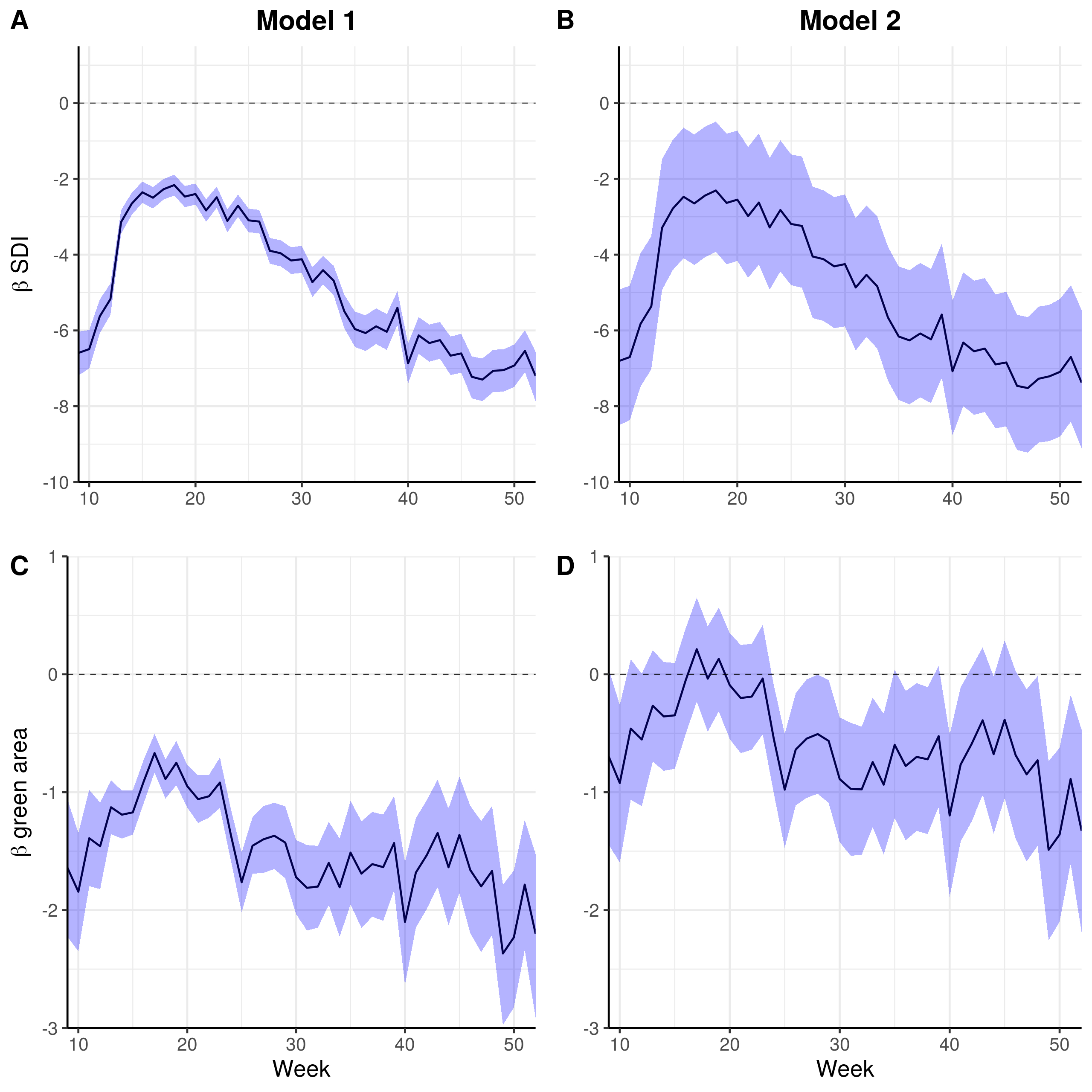

### S3 Fig

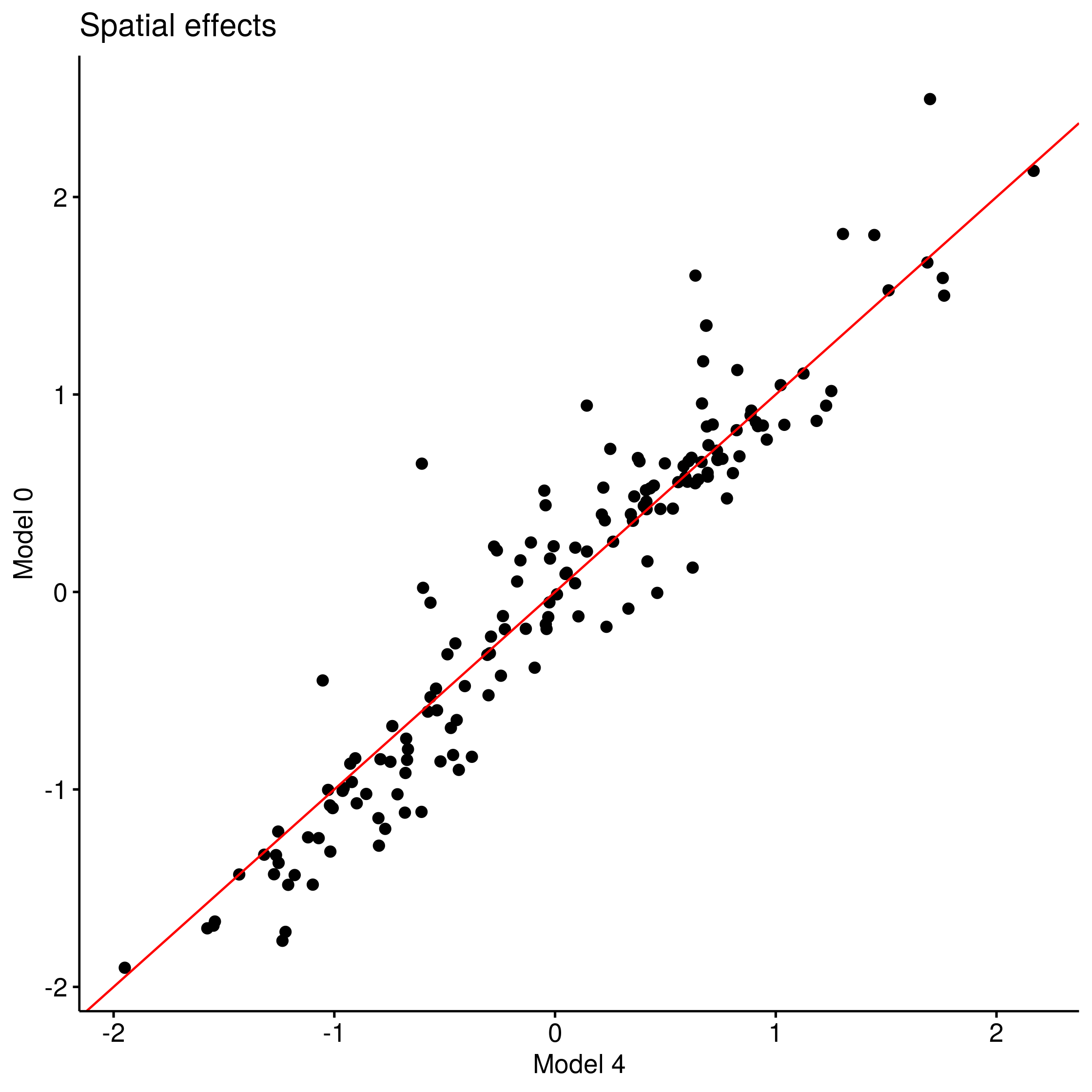

### S4 Fig

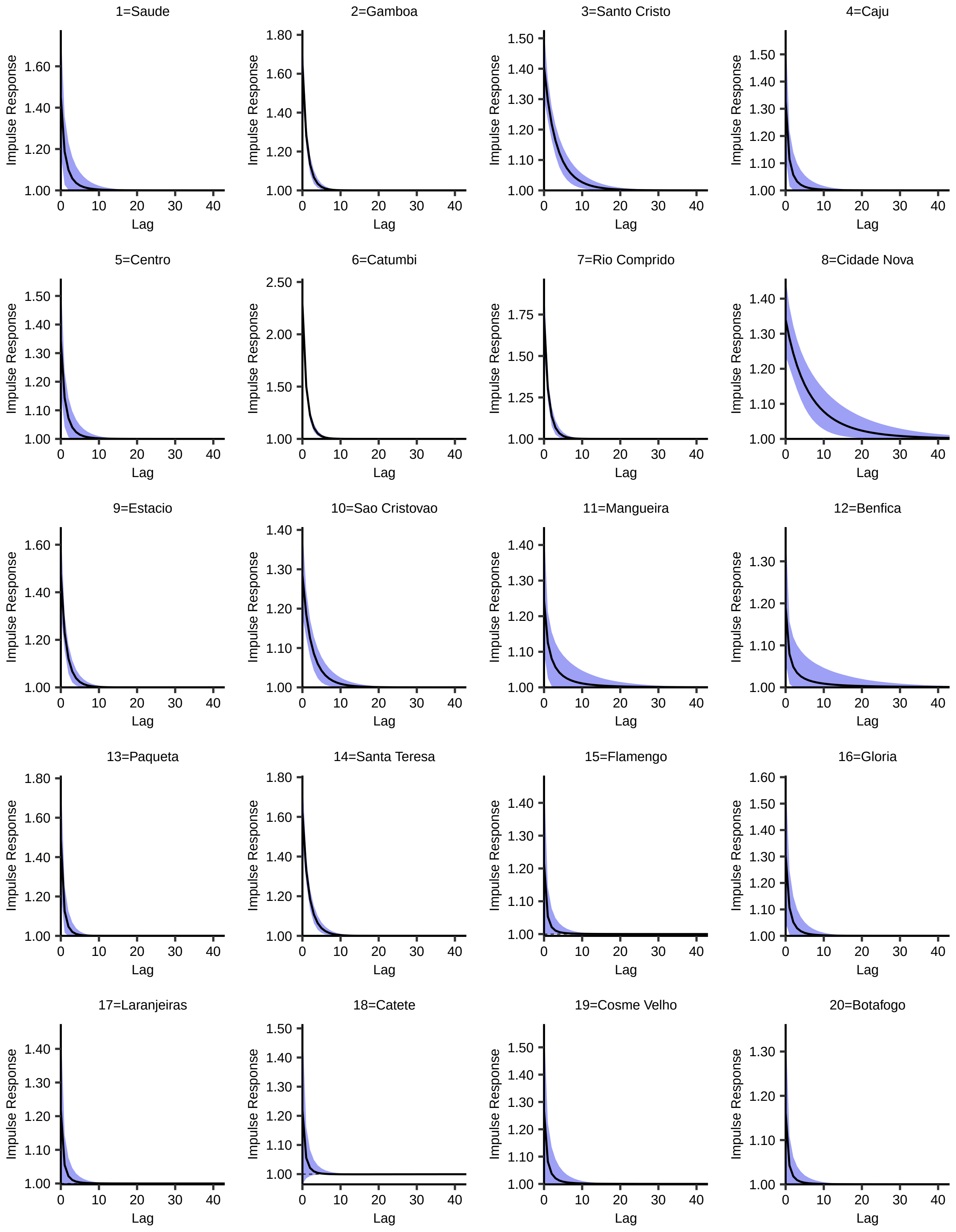

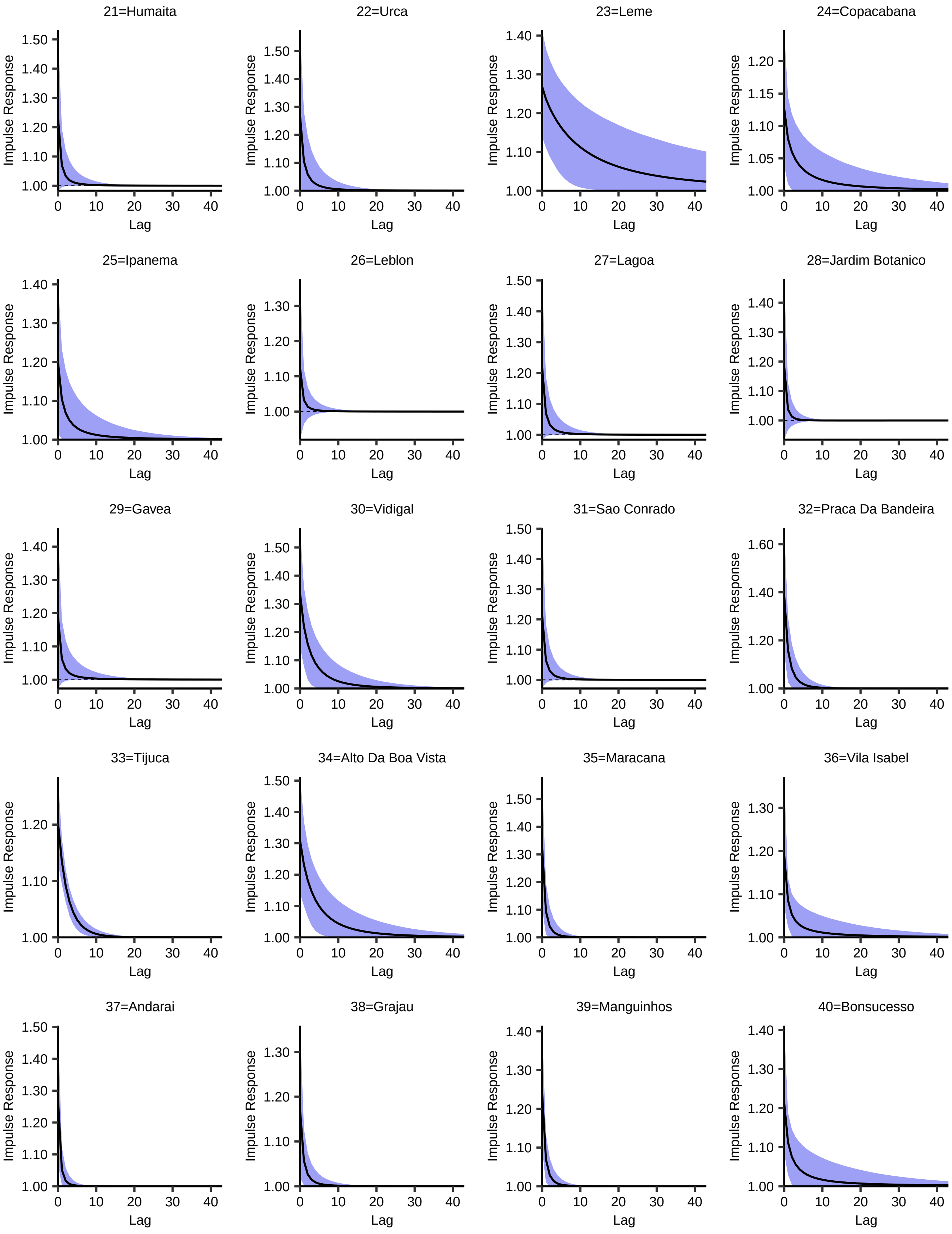

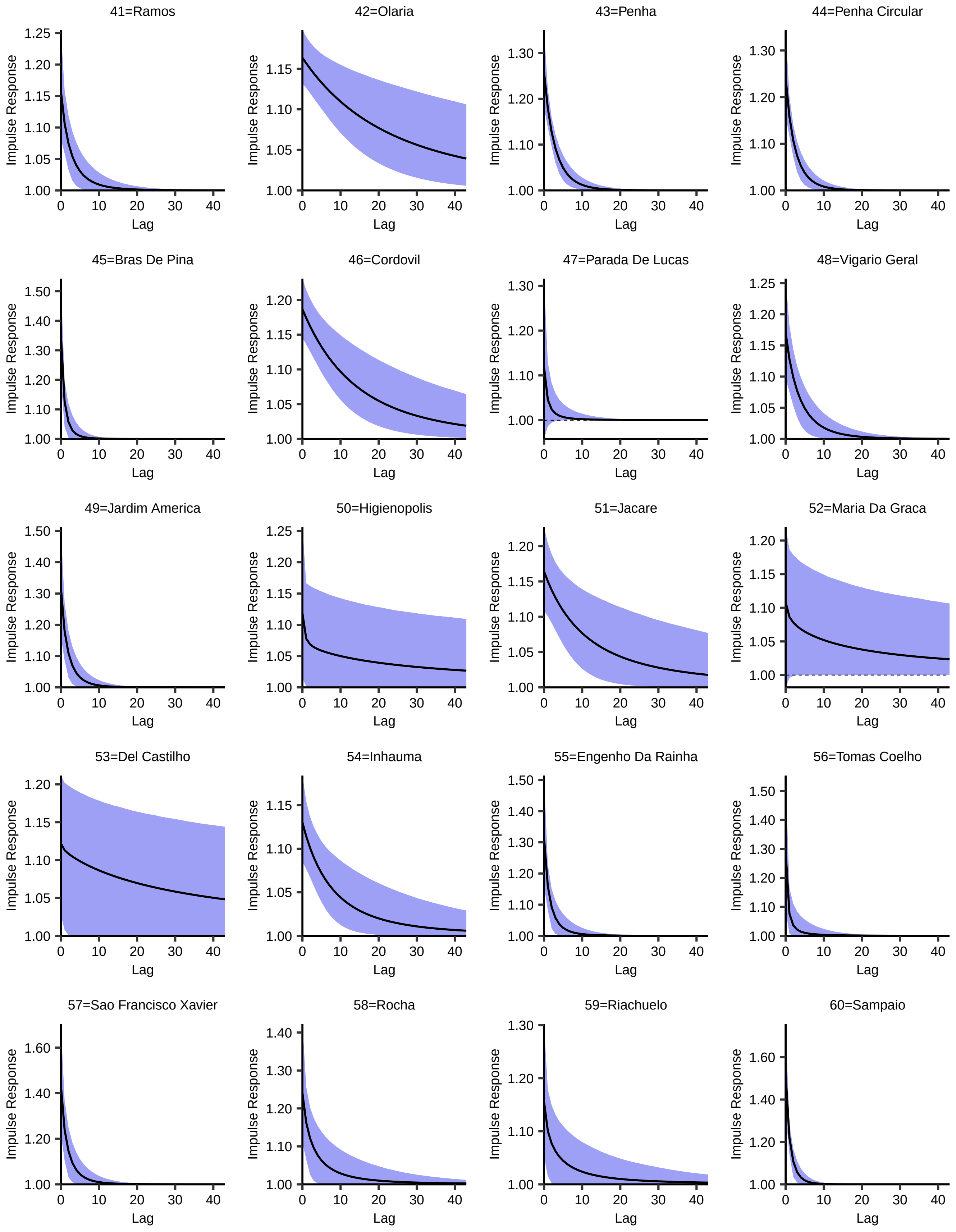

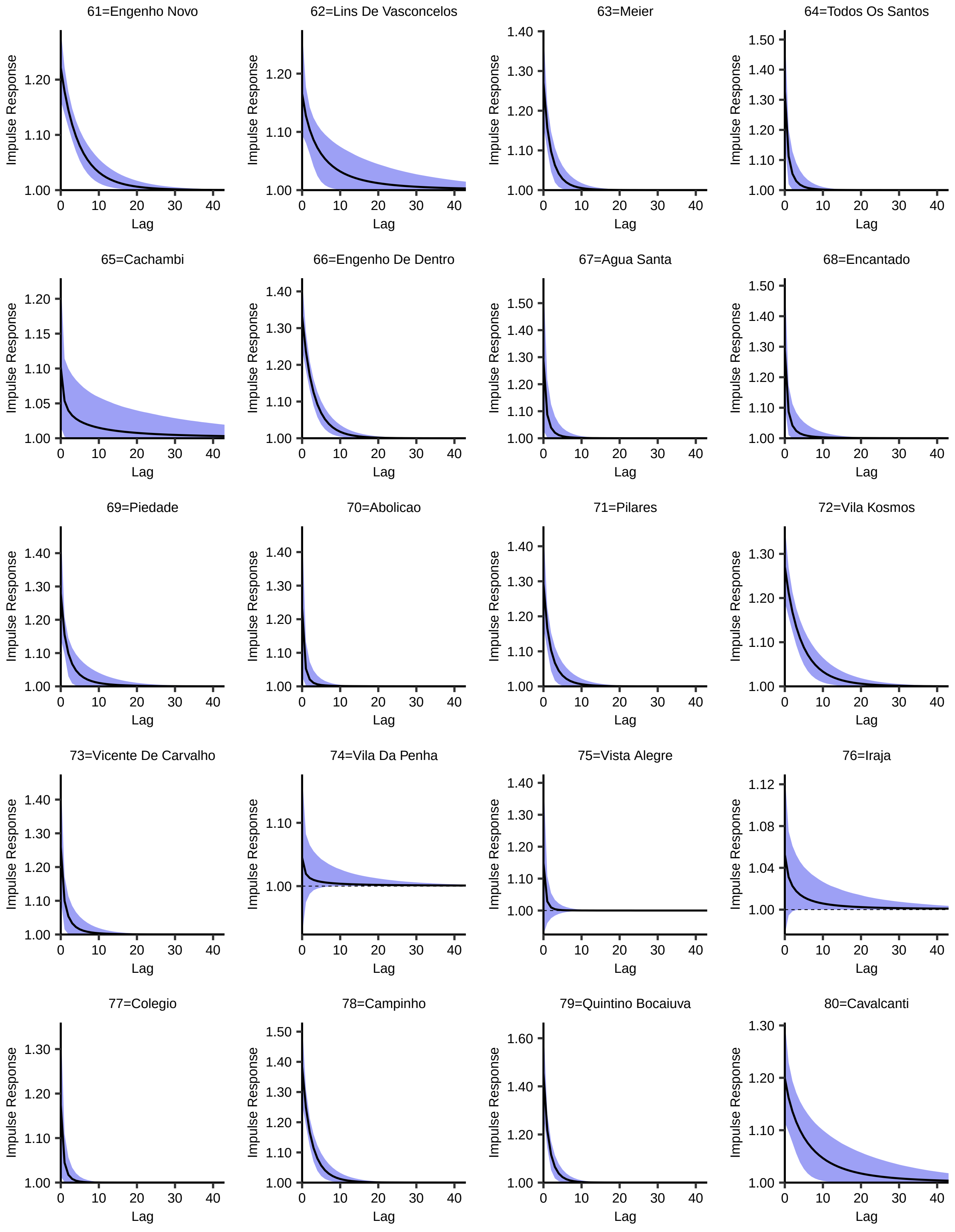

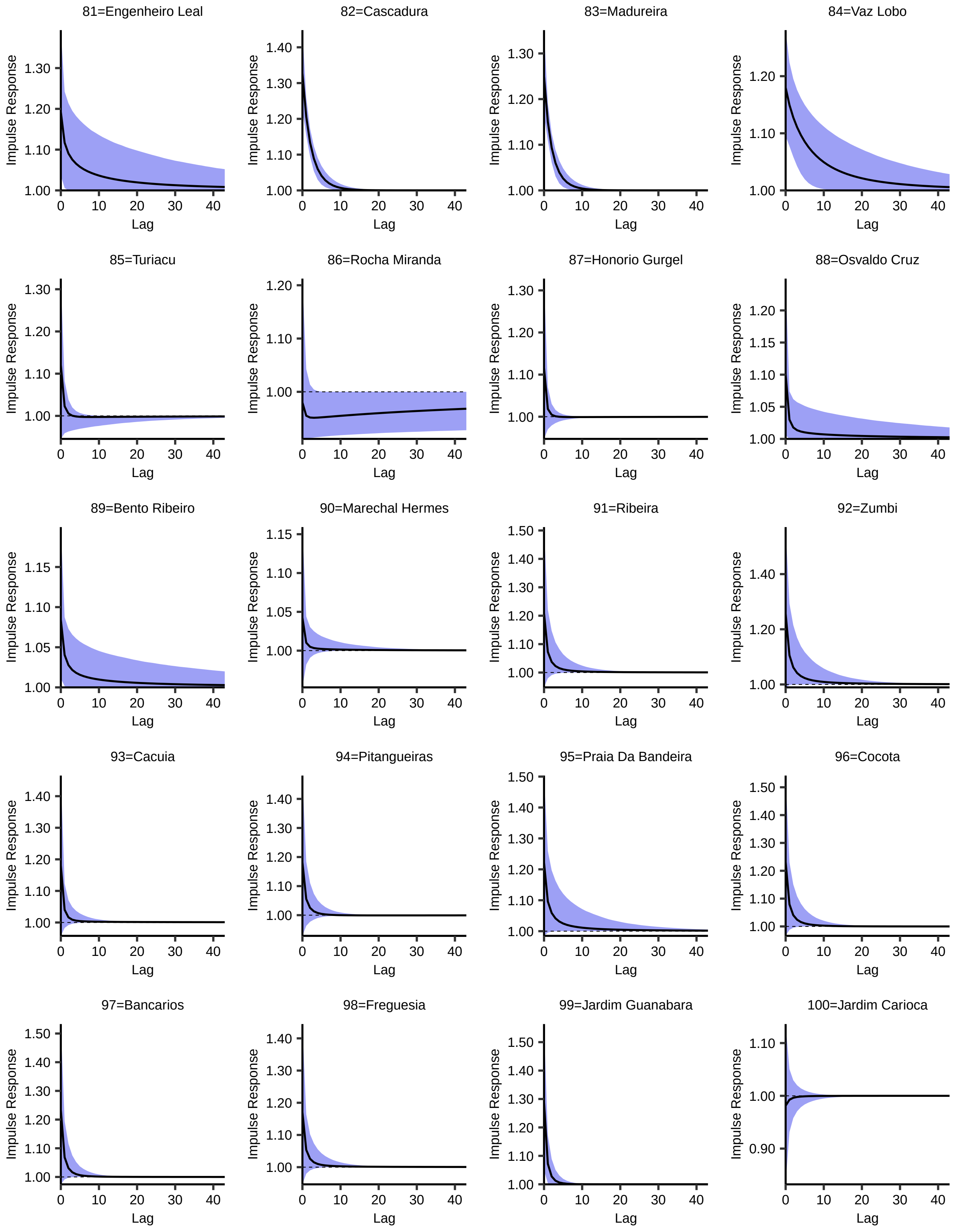

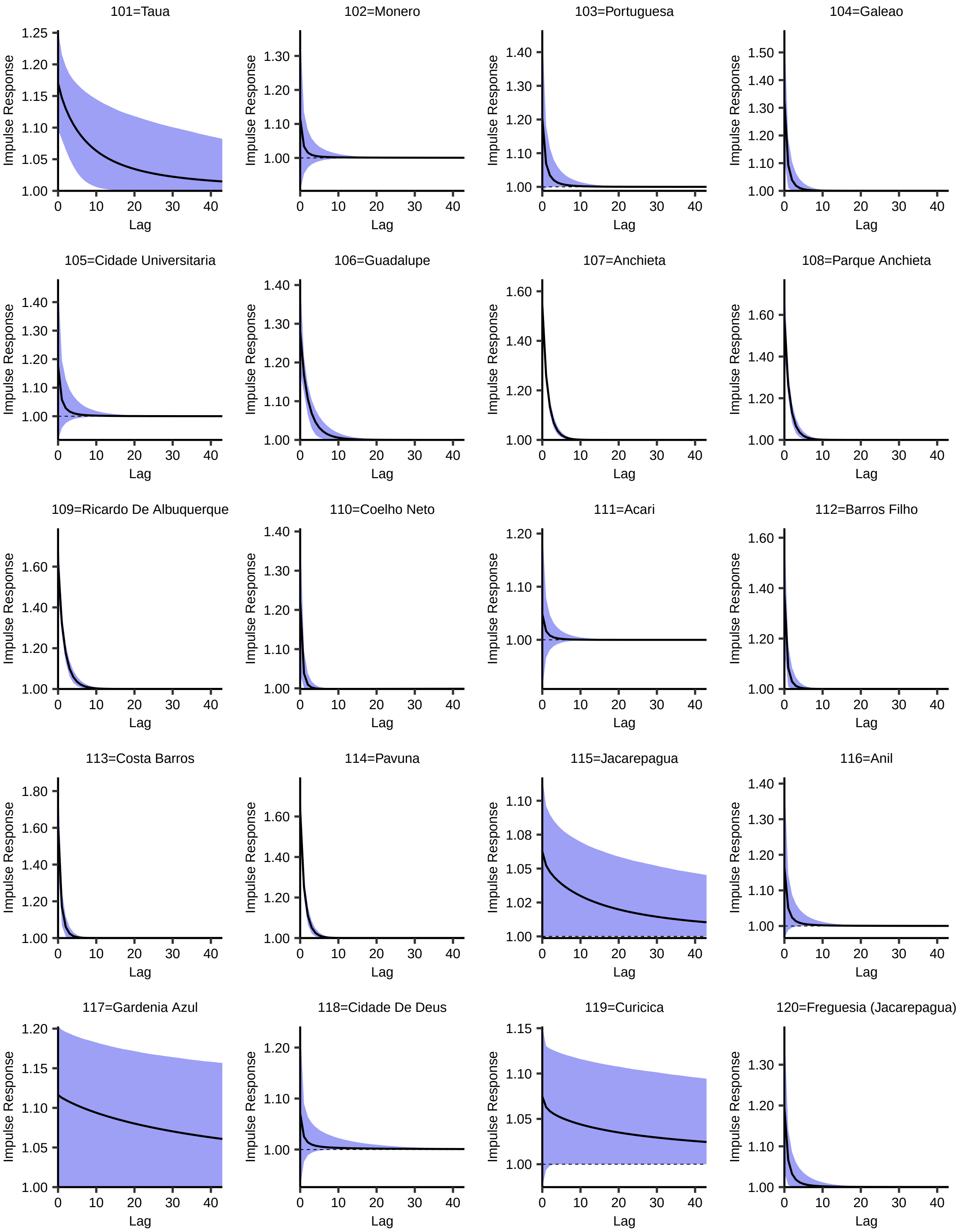

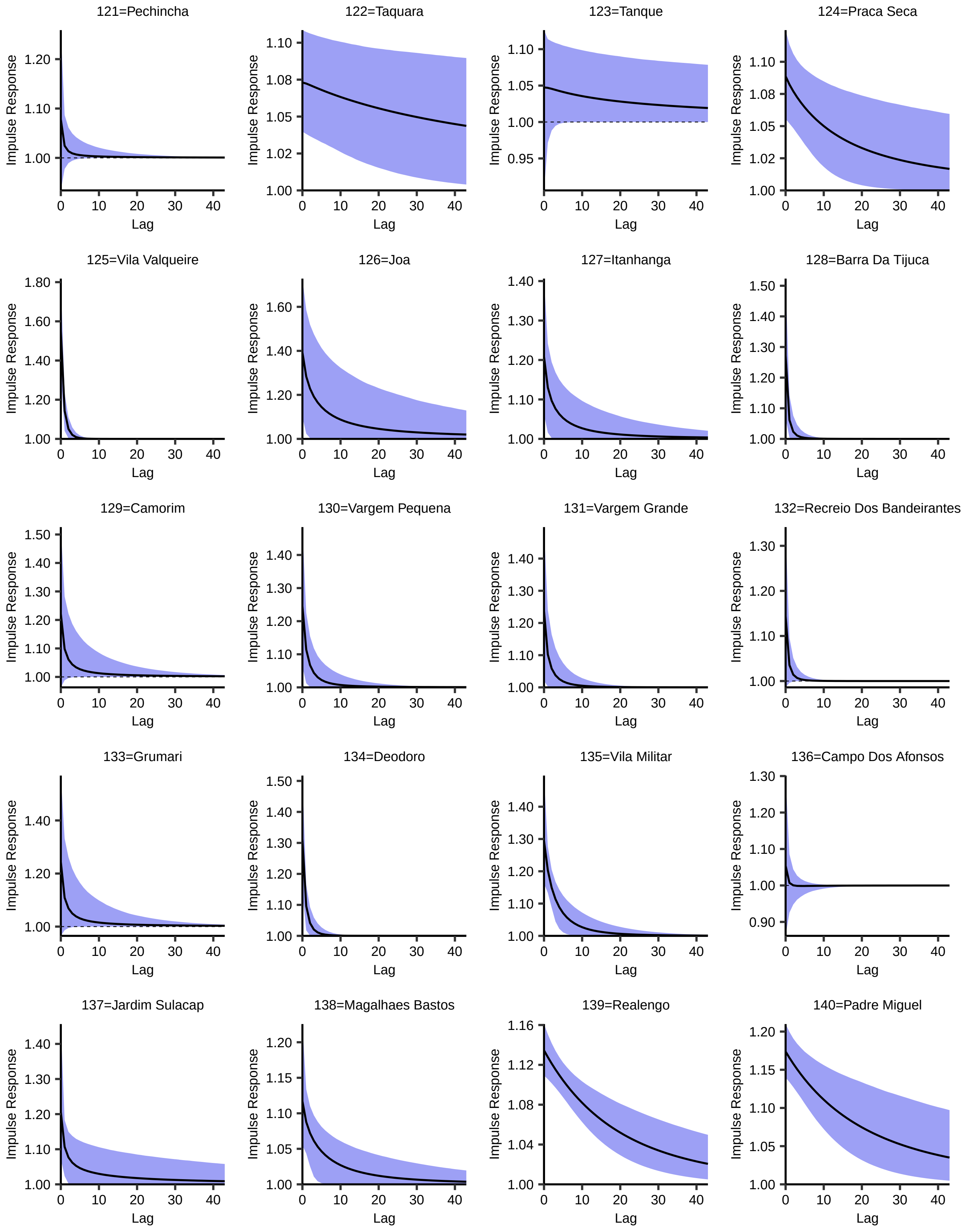

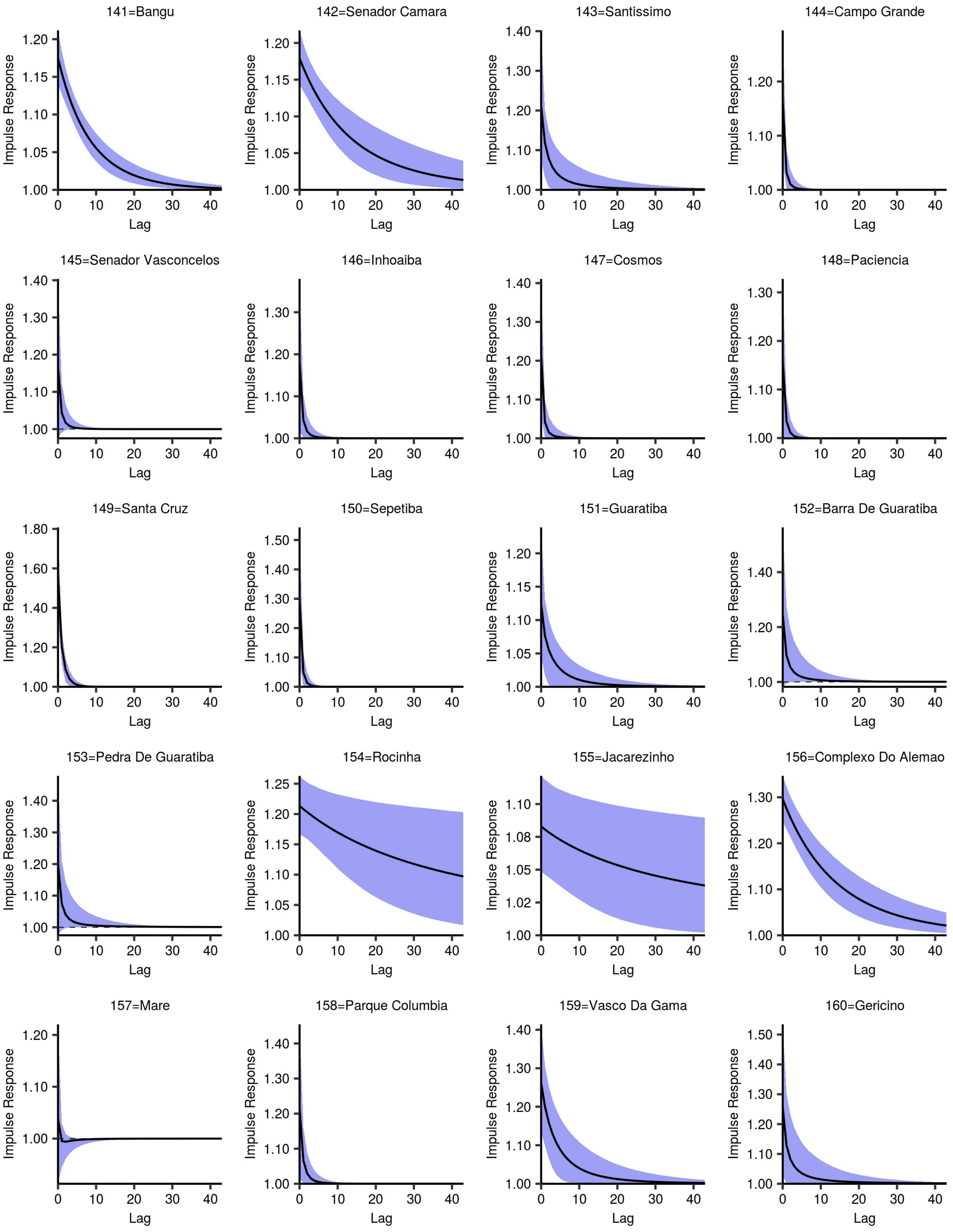

### S5 Fig

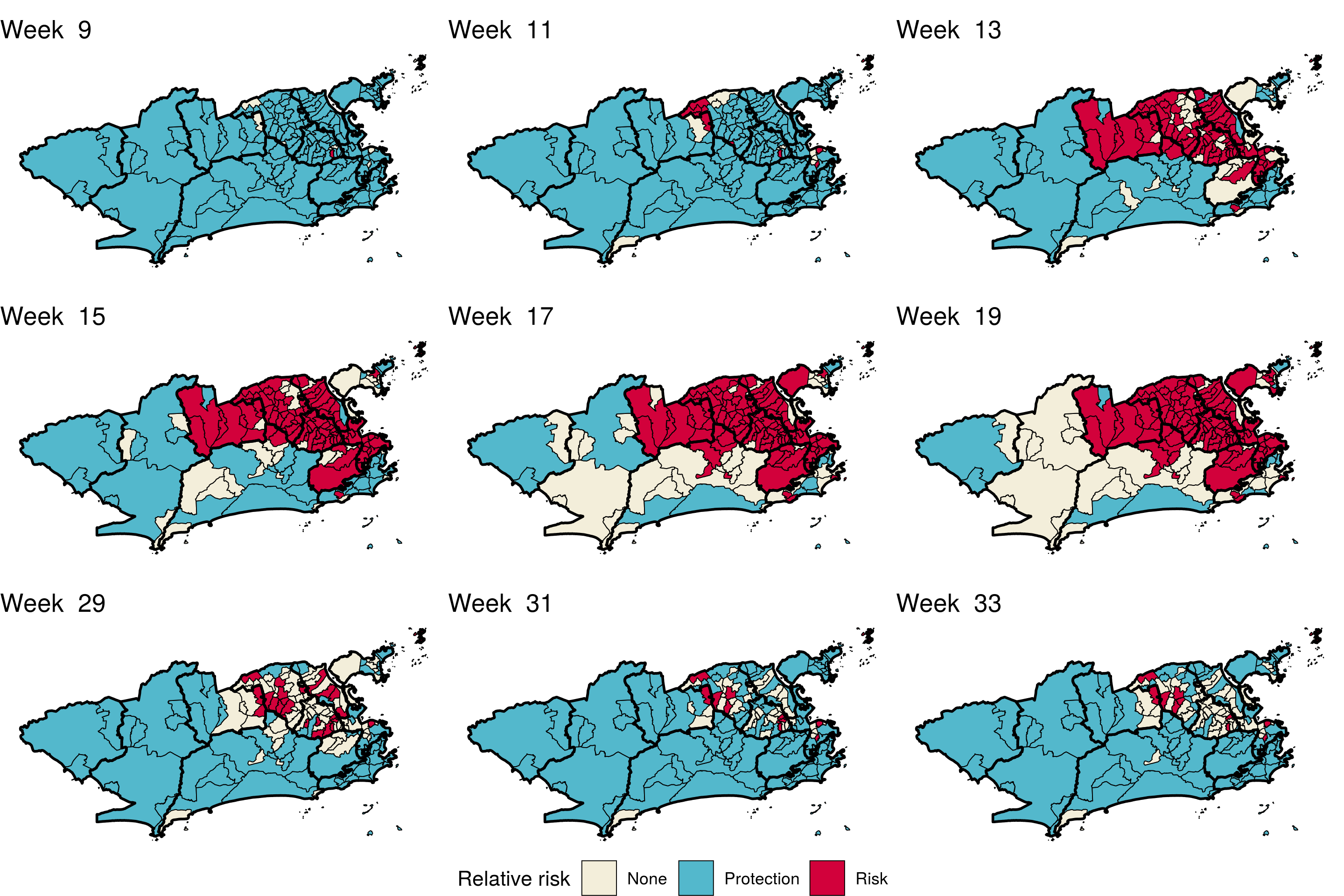
